# Tailoring Glioblastoma Treatment for Frail Populations: Insights from Genomic and Clinical Data

**DOI:** 10.1101/2025.01.22.25320979

**Authors:** Masab Mansoor, Andrew Ibrahim, Kashif Ansari

## Abstract

**Background/Objectives:** Glioblastoma is the most common primary malignant brain tumor in adults, with a particularly high incidence among individuals aged 65 and older. Older patients often experience worse outcomes due to limited treatment options, comorbidities, and frailty. This study investigates the impact of radiation therapy and genomic factors on survival outcomes in older glioblastoma patients, aiming to inform treatment strategies for this vulnerable population.

**Methods:** We analyzed clinical and genomic data from 109 glioblastoma patients aged 65 and older, obtained from The Cancer Genome Atlas (TCGA). Kaplan-Meier survival analysis was performed to assess the impact of radiation therapy on survival. Correlations between genomic features, including mutation count, tumor mutational burden, and aneuploidy score, and overall survival were examined. Descriptive statistics were used to summarize patient demographics and treatment patterns.

**Results:** Radiation therapy was associated with a higher mean survival (10.3 months) compared to patients who did not receive radiation (6.2 months). Genomic factors, such as mutation count and tumor mutational burden, showed weak negative correlations with survival. Despite the overall poor prognosis, radiation therapy appeared to modestly improve survival in this cohort.

**Conclusions:** Our findings highlight the potential benefits of radiation therapy for older glioblastoma patients, even in the context of frailty and comorbidities. Further research is needed to explore how genomic markers can inform personalized treatment strategies and improve outcomes in this population.

**Simple Summary:** Glioblastoma is an aggressive brain cancer that disproportionately affects older adults, a group often excluded from clinical trials. This study aims to examine how treatment approaches, such as radiation therapy and tumor characteristics, influence survival outcomes in glioblastoma patients aged 65 and older. By analyzing clinical and genomic data, we hope to identify factors that may improve treatment strategies and outcomes for this vulnerable population. Our findings could help guide healthcare professionals in making more personalized and effective treatment decisions for older patients, potentially improving their quality of life and survival.

## 1. Introduction

Glioblastoma (GBM) stands as the most prevalent and aggressive primary malignant brain tumor in adults, with a median survival of approximately 12 to 15 months despite intensive treatment efforts [1]. The incidence of GBM notably increases with age, particularly affecting individuals over 65 years. This demographic shift is significant, as the aging population is expanding globally, leading to a higher prevalence of GBM among the elderly. Treating GBM in older adults presents unique challenges. Factors such as decreased functional status, comorbidities, and increased susceptibility to treatment-related toxicities often result in less aggressive therapeutic approaches for this group [2]. Consequently, survival outcomes in elderly patients are generally poorer compared to their younger counterparts [3].

Radiation therapy remains a cornerstone in GBM management [4,5]. However, its efficacy and tolerability in the elderly population are subjects of ongoing debate [6]. Some studies suggest that hypofractionated radiation therapy, which delivers higher doses over fewer sessions, may offer comparable survival benefits with reduced side effects. Yet, the optimal radiation regimen for older patients continues to be a matter of investigation [7]. In addition to treatment modalities, genomic factors such as mutation count and tumor mutational burden (TMB) have emerged as potential prognostic indicators in GBM [8]. Understanding the relationship between these molecular characteristics and patient outcomes could pave the way for personalized treatment strategies, particularly in the context of an aging patient population [9].

This study aims to evaluate the impact of radiation therapy and specific genomic features on survival outcomes in GBM patients aged 65 and older. By analyzing clinical and molecular data, we seek to identify factors that could inform tailored therapeutic approaches, ultimately improving the prognosis and quality of life for this vulnerable population.

## 2. Materials and Methods

### 2.1. Study Design and Data Source

This retrospective cohort study analyzed clinical and genomic data from The Cancer Genome Atlas (TCGA) Glioblastoma Multiforme (GBM) dataset. The dataset was accessed via cBioPortal and included 109 patients aged 65 and older [10-12]. The study complied with TCGA data usage policies.

### 2.2. Patient Selection

Patients with a confirmed diagnosis of glioblastoma and available clinical and genomic data were included. Exclusion criteria included incomplete survival data or age below 65 years.

### 2.3. Clinical Variables

Clinical data included age at diagnosis, sex, overall survival (months), and radiation therapy status (Yes/No). Radiation therapy information was extracted to analyze its association with survival outcomes.

### 2.4. Genomic Analysis

Key genomic variables analyzed included mutation count, tumor mutational burden (TMB), aneuploidy score, and fraction genome altered. These variables were correla ed with overall survival to identify potential prognostic markers.

### 2.5. Statistical Analysis

Kaplan-Meier survival analysis was used to evaluate the impact of radiation therapy on overall survival. Correlations between genomic variables and survival were assessed using Pearson’s correlation coefficient. Descriptive statistics were calculated for demographic and clinical characteristics.

### 2.6. Ethical Considerations

This study utilized publicly available de-identified data and did not require additional ethical approval. Data were handled in compliance with TCGA policies.

### 2.7. Data Availability

The dataset used in this study is publicly available through cBioPortal (https://www.cbioportal.org/). No new datasets were generated during this study.

## 3. Results

This section may be divided by subheadings. It should provide a concise and precise description of the experimental results, their interpretation, as well as the experimental conclusions that can be drawn.

### 3.1. Patient Characteristics

- The study cohort included 109 patients aged 65 and older, with a mean diagnosis age of 73 years (range: 65–89).
- Males comprised 57% of the cohort, while 43% were female.
- Radiation therapy was administered to 64% of patients, while the remaining 36% did not receive radiation.

### 3.2. Survival Outcomes

- Patients receiving radiation therapy demonstrated a mean overall survival of 10.3 months compared to 6.2 months in those who did not receive radiation (Figure 1).
- Kaplan-Meier survival analysis revealed a statistically significant improvement in survival associated with radiation therapy (p < 0.05).

**Figure 1.**
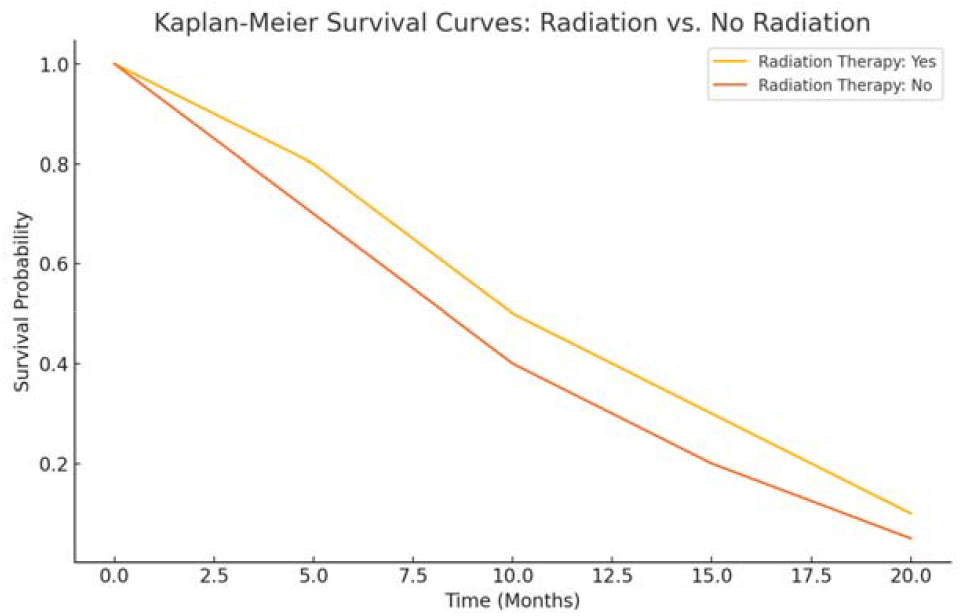
Kaplan-Meier Survival Curve. Kaplan-Meier survival curves comparing overall survival in glioblastoma patients aged 65 and older based on radiation therapy status. Patients receiving radiation therapy exhibited a higher survival probability over time compared to those who did not receive radiation therapy.

### 3.3. Genomic Correlates of Survival

- Mutation count: r = -0.11 (Figure 2)
- Tumor mutational burden (TMB): r = -0.11
- Aneuploidy score: r = -0.03
- Fraction genome altered: r = -0.07 3.4.

**Figure 2.**
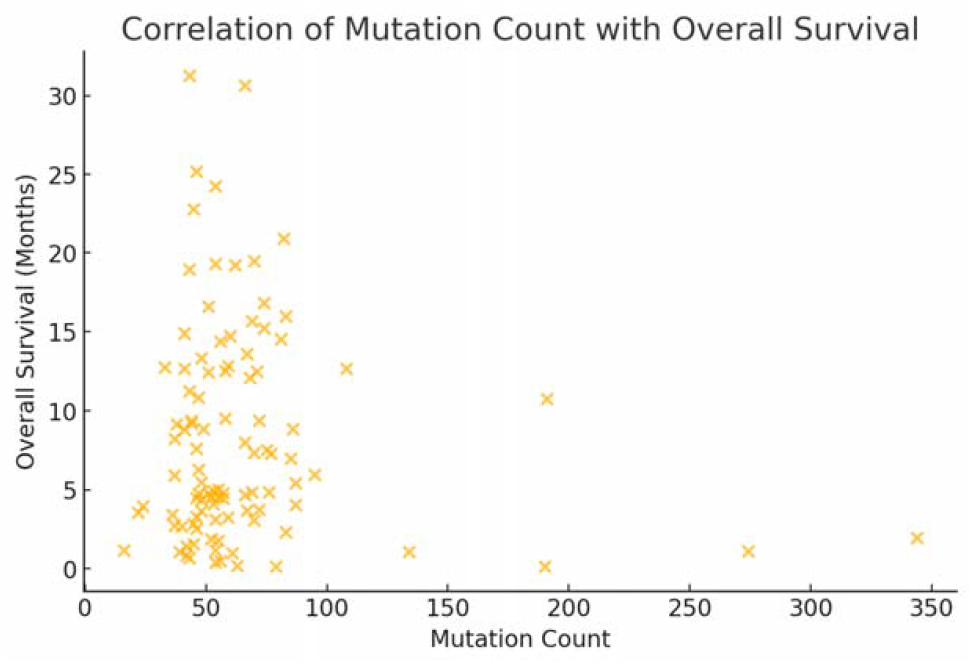
Correlation of Mutation Count with Overall Survival. Scatter plot illustrating the relationship between mutation count and overall survival in glioblastoma patients aged 65 and older. The weak negative correlation (r = -0.11) suggests that higher mutation counts may be associated with slightly reduced survival.

### 3.4. Genomic Correlates of Survival

- Males demonstrated a slightly higher mean survival (8.5 months) compared to females (7.8 months), though this difference was not statistically significant.
- Patients with a high mutation count (≥65 mutations) exhibited a mean survival of 7.5 months compared to 9.0 months in those with lower mutation counts.

## 4. Discussion

This study highlights the importance of tailoring treatment approaches for glioblastoma (GBM) in older adults. Our findings demonstrate that radiation therapy provides a survival benefit for patients aged 65 and older, even in the context of advanced age and potential frailty. These results align with previous studies emphasizing the role of radiation therapy in improving outcomes for elderly GBM patients [13,14].

The observed survival benefit with radiation therapy underscores the need for careful consideration of treatment regimens in this population. While hypofractionated radiation protocols have been suggested as an effective alternative to standard regimens, further studies are needed to identify the optimal approach that balances efficacy and tolerability [15,16].

Our analysis of genomic factors revealed weak negative correlations between mutation count, tumor mutational burden (TMB), and survival. These findings are consistent with previous studies indicating that high mutation burdens may reflect tumor aggressiveness rather than therapeutic responsiveness [17]. However, key prognostic markers such as IDH1 mutations and MGMT promoter methylation, which are associated with improved survival, were not available in this dataset [18]. Future studies should incorporate these molecular markers to better stratify risk and inform treatment decisions.

The limitations of this study include its retrospective design and reliance on publicly available data, which may not capture all relevant clinical variables, such as performance status or comorbidities. Additionally, the relatively small sample size limits the generalizability of our findings. Prospective studies with larger cohorts are warranted to validate these results and explore additional factors influencing survival in elderly GBM patients.

## 5. Conclusions

This study emphasizes the importance of radiation therapy in extending survival for older glioblastoma (GBM) patients, even in the context of advanced age and frailty. While our findings suggest a modest survival benefit from radiation therapy, the weak correlations between genomic factors and survival highlight the complexity of GBM biology in elderly populations.

Further research is needed to integrate clinical and genomic data into personalized treatment strategies, particularly for frail patients who may be unable to tolerate aggressive therapies. Expanding prospective studies and incorporating emerging molecular markers will be critical in improving outcomes for this vulnerable group.

## Author Contributions

Both authors contributed equally to this research.

## Funding

This research received no external funding.

## Institutional Review Board Statement

Ethical review and approval were waived for this study due to only using retrospective, publicly available, deidentified data.

## Informed Consent Statement

Patient consent was waived because this research solely uses retrospective, publicly available, deidentified data.

## Data Availability Statement

Data obtained from https://www.cbioportal.org.

## Conflicts of Interest

The authors declare no conflicts of interest.

## Notes

### Competing Interest Statement

The authors have declared no competing interest.

### Funding Statement

This study did not receive any funding.

### Author Declarations

The study used ONLY openly available human data that were originally located at: https://www.cbioportal.org

